# Association of 25 hydroxyvitamin D concentration with risk of COVID-19: a Mendelian randomization study

**DOI:** 10.1101/2020.08.09.20171280

**Authors:** Di Liu, Qiuyue Tian, Jie Zhang, Haifeng Hou, Wei Wang, Qun Meng, Youxin Wang

**Affiliations:** Beijing Key Laboratory of Clinical Epidemiology, School of Public Health, Capital Medical University, Beijing, 100069, China; School of Public Health, Shandong First Medical University & Shandong Academy of Medical Sciences, Tai’an 271016, Shandong Province, China; School of Medical and Health Sciences, Edith Cowan University, Perth 60127, Australia; Inner Mongolia Comprehensive Center for Disease Control and Prevention, Hohhot, Inner Mongolia Autonomous Region, China

**Keywords:** Coronavirus disease 2019, vitamin D deficiency, 25 hydroxyvitamin D, Mendelian randomization

## Abstract

**Background:** In observational studies, 25 hydroxyvitamin D (25OHD) concentration has been associated with an increased risk of Coronavirus disease 2019 (COVID-19). However, it remains unclear whether this association is causal.

**Methods:** We performed a two-sample Mendelian randomization (MR) to explore the causal relationship between 25OHD concentration and COVID-19, using summary data from the genome-wide association studies (GWASs) and using 25OHD concentration-related SNPs as instrumental variables (IVs).

**Results:** MR analysis did not show any evidence of a causal association of 25OHD concentration with COVID-19 susceptibility and severity (odd ratio [OR]=1.136, 95% confidence interval [CI] 0.988-1.306; *P*=0.074 OR=0.889, 95% CI 0.549-1.439 *P*=0.632). Sensitivity analyses using different instruments and statistical models yielded similar findings, suggesting the robustness of the causal association. No obvious pleiotropy bias and heterogeneity were observed.

**Conclusion:** The MR analysis showed that there might be no linear causal relationship of 25OHD concentration with COVID-19 susceptibility and severity.

## Introduction

Coronavirus disease 2019 (COVID-19) has become a global pandemic ^1^. Aimed at delaying disease onset by modulating modifiable risk factors, primary prevention has been proposed as a potentially effective and feasible tool to address the global challenge posed by COVID-19.

Vitamin D is a hormone, produced in the skin during exposure to sunlight, helped regulate the amount of calcium and phosphate in the body, which are needed to keep bones, teeth, and muscles healthy, and played a critical role in the immune system ^2-4^. Vitamin D deficiency is a candidate risk factor for a range of adverse health outcomes, such as cancer, cardiovascular diseases, metabolic disorders, infectious diseases, as well as autoimmune diseases ^5,6^. There were well recognized evidences that vitamin D deficiency contributes to the seasonal increase of virus infections of the respiratory tract, from the common cold to influenza, and now possibly also COVID-19 ^7,8^. Some systematic reviews indicated that vitamin D deficiency may increase infection risk of COVID-19 by discussing the biological mechanism of virus infection^4,9,10^. In addition, numerous population-based studies have evaluated the Vitamin D deficiency in COVID-19 patients relative to controls; however, the findings have been inconsistent. Some studies showed that COVID-19 patients had a lower 25 hydroxyvitamin D (25OHD) concentration compared with healthy controls ^7,11-13^, while others found no association ^14-16^. The inconsistent findings from these epidemiological studies may be due to differences in the study design, study population, assays used for measuring Vitamin D deficiency. It is important to note that as the nature of the above studies is observational, such traditional epidemiological studies are particularly vulnerable to reverse causality and residual confounding.

A promising approach known as Mendelian randomization (MR), which uses inherited genetic variants as instrumental variables, provides stronger evidence for the causal effect of exposure on the diseases largely overcoming the traditional limitations due to confounding and reverse causality ^17,18^. Summary based MR is an excellent strategy to evaluate the causality using summary statistics from Genome Wide Association Study (GWAS) data^19,20^. Therefore, we conducted MR analysis to assess whether 25OHD concentration was causally associated with risk of COVID-19.

## Methods

### Data sources

#### Genetic association datasets for COVID-19 susceptibility and severity

The summarized data was obtained from the most recent version of GWAS analyses from the COVID-19 host genetics initiative from UK Biobank individuals, which released on July 1, 2020 (https://www.covid19hg.org/results/)^21^. Summarized data on COVID-19 included two population, COVID-19 (n=6,696) vs. general population without the phenotype (n=1073072) and COVID-19 (n=3,523) vs. COVID-19 negative (n=36,634); and summarized data on severe COVID-19 included 536 patients and 329391 control participants from general population without the phenotype. In addition, we used summarized data for severe respiratory confirmed COVID-19 reported by from the Severe Covid-19 GWAS Group^22^. The GWAS summarized data can be downloaded at https://ikmb.shinyapps.io/COVID-19_GWAS_Browser/. The study included 1,160 patients who had severe respiratory confirmed COVID-19, and 2,205 participants from the general population without COVID-19 as the control.

#### Selection of 25OHD concentration -associated single nucleotide polymorphisms (SNPs)

We selected 143 SNPs as instrumental variables (IVs) for 25OHD concentration based on the recent large-scale GWAS ^23^ In addition, we retained independent variants from each other (Linkage disequilibrium [LD], r < 0.001) for sensitivity analysis. When we encountered genetic variants in LD, the SNP with the lowest *p*-value was selected. The LD proxies were defined using 1000 genomes European samples.

### MR analysis

In the analyses, the inverse-variance weighted (IVW) method was used to estimate the overall causal association of 25OHD concentration on COVID-19 susceptibility and severity. We additionally conducted the weighted median, penalized weighted median, and MR-Egger regression to account for potential violations of valid instrumental variable assumptions. The MR-Egger analysis was performed to evaluate pleiotropy based on the intercept. We conducted a heterogeneity test in MR analyses using the IVW Q test. Then, sensitivity analyses were performed to examine the stability of the causal estimate. Firstly, we performed a “leave one out” analysis to further investigate the possibility that the causal association was driven by a single SNP. Then, we retained independent variants from each other (LD, r^2^ < 0.001) for further sensitivity analysis. Results were presented as odds ratios (OR) with their 95% confidence interval (CI) and beta with standard error (SE) of outcomes per genetically predicted increase in each exposure factor.

In terms of various estimates for different measures, we chose the result of main MR method as the following rules:

1. If no directional pleiotropy in MR estimates (Q statistic: *P* value > 0.05, MR-Egger intercept: *P* value > 0.05), the results of the IVW method were reported.
2. If directional pleiotropy was detected (MR-Egger intercept: *P* value < 0.05) and *P* value > 0.05 for the Q test, the results of the MR-Egger method were reported.
3. If directional pleiotropy was detected (MR-Egger intercept: *P* value < 0.05) and *P* value < 0.05 for the Q test, the results of the weighted median method were reported.

All data analyses were performed by the “twosampleMR” package using R version 4.0.0 (https://www.r-project.org/).

## Results

As shown in **Table 1**, the MR analysis showed no significant association of genetically instrumented 25OHD concentration with COVID-19 in the population of COVID-19 vs. population and COVID-19 vs. COVID-19 negative (OR=1.136, 95% CI 0.988-1.306, *P*=0.074; OR=1.168, 95% CI 0.956-1.427, *P*=0.128). The association of 25OHD concentration with COVID-19 was robust in the weighted median and penalized weighted median methods, except in the MR-Egger regression (OR, 1.258; 95% CI, 1.053-1.502; *P*=0.013; OR, 1.302; 95% CI, 1.011-1.676; *P*=0.044). Pleiotropy bias and heterogeneity were also not observed. In terms of various estimates for different measures, we reported the results of the IVW method. In addition, the “leave one out” results showed that by omitting the included 89 SNPs one at a time, no individual genetic variants seem to have any significant effect on the overall results (**Figure 1-2**). The association of 25OHD concentration with COVID-19 remained robust using SNP instrument (LD, r^2^ < 0.001) (**Table 1, Figure S1**).

**Table 1.** Causal association of 25OHD concentration with COVID-19.

| Method | SNP (n) | OR (95% CI) | Beta (SE) | *P* | SNP (n) | OR (95% CI) | Beta (SE) | *P* |
| --- | --- | --- | --- | --- | --- | --- | --- | --- |
| COVID-19 vs. general population |  |  |  |  |  |  |  |  |
| IVW | 89 | 1.136 (0.988-1.306) | 0.128 (0.071) | 0.074 | 56 | 1.048 (0.882-1.245) | 0.047 (0.088) | 0.596 |
| Weighted median | 89 | 1.155 (0.936-1.425) | 0.144 (0.107) | 0.179 | 56 | 1.142 (0.915-1.426) | 0.133 (0.113) | 0.239 |
| Penalised weighted median | 89 | 1.155 (0.939-1.420) | 0.144 (0.106) | 0.173 | 56 | 1.142 (0.913-1.429) | 0.133 (0.114) | 0.244 |
| MR_Egger | 89 | 1.258 (1.053-1.502) | 0.229 (0.090) | 0.013 | 56 | 1.149 (0.918-1.439) | 0.139 (0.115) | 0.230 |
| $\beta$ (intercept) | 89 | - | -0.007 (0.004 ) | 0.071 | 56 | - | -0.006 (0.005) | 0.214 |
| Q statistic | 89 | - | - | 0.759 | 56 | - | - | 0.794 |
| COVID-19 vs. COVID-19 negative |  |  |  |  |  |  |  |  |
| IVW | 89 | 1.168 (0.956-1.427) | 0.156 (0.102) | 0.128 | 57 | 1.192 (0.945-1.504) | 0.176 (0.118) | 0.138 |
| Weighted median | 89 | 1.017 (0.750-1.379) | 0.017 (0.156) | 0.912 | 57 | 1.134 (0.810-1.588) | 0.126 (0.172) | 0.464 |
| Penalised weighted median | 89 | 0.990 (0.737-1.328) | -0.010 (0.150) | 0.945 | 57 | 1.134 (0.814-1.579) | 0.126 (0.169) | 0.4570. |
| MR_Egger | 89 | 1.302 (1.011-1.676) | 0.26 (0.13) | 0.044 | 57 | 1.265 (0.951-1.682) | 0.235 (0.145) | 0.112 |
| $\beta$ (intercept) | 89 | - | -0.007 (0.005) | 0.174 | 57 | - | -0.004 (0.006) | 0.485 |
| Q statistic | 89 | - | - | 0.656 | 57 | - | - | 0.925 |

Beta is the estimated effect size.
25OHD: 25 hydroxyvitamin D; CI: confidence intervals; COVID-19: Coronavirus disease 2019; IVs: instrumental variables; IVW, inverse-variance weighted; MR mendelian randomization; OR: odds ratio; SE, standard error; SNP, single-nucleotide polymorphism.

**Figure 1.**
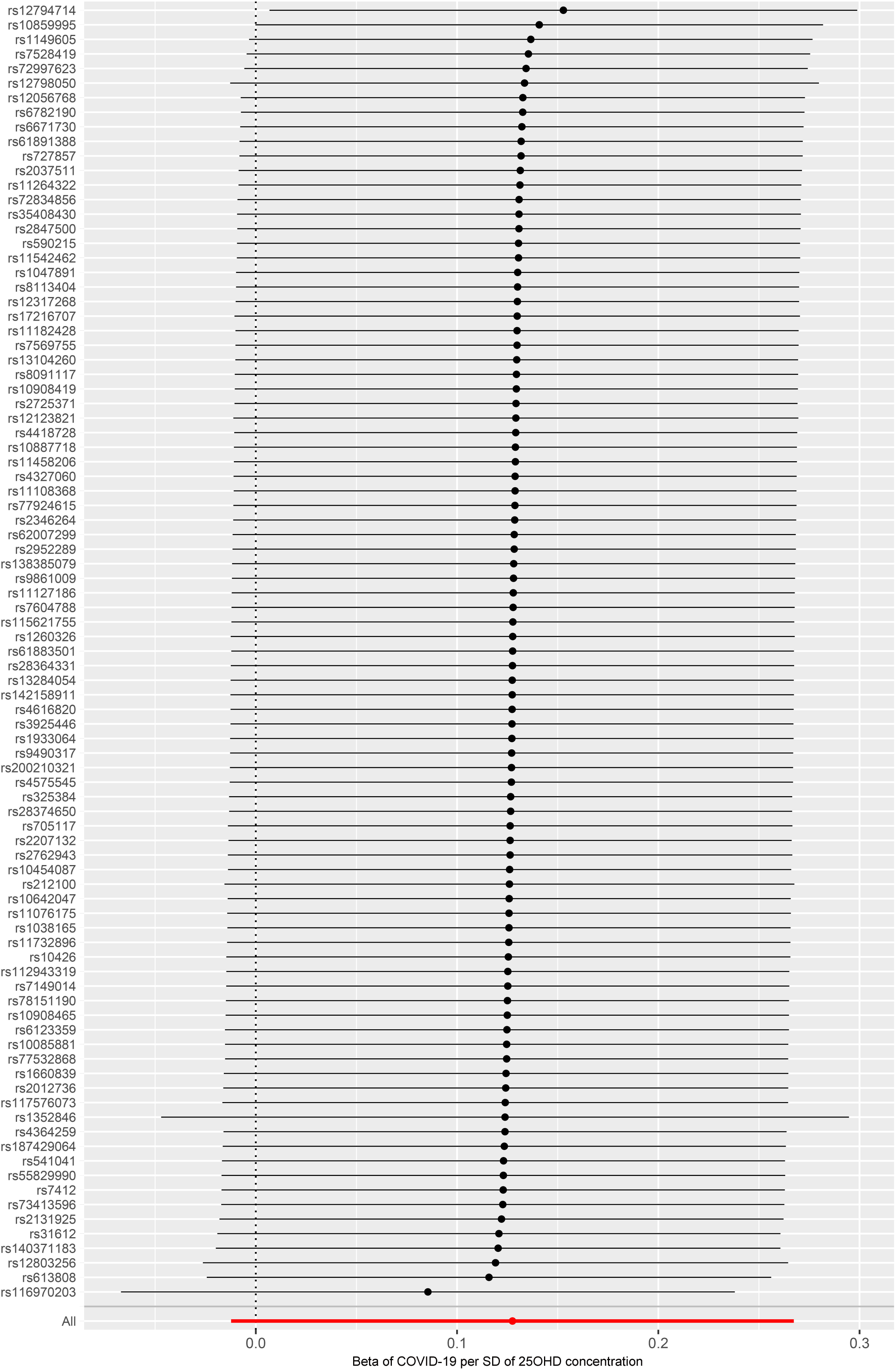
MR leave-one-out sensitivity analysis for ‘25OHD concentration’ on ‘COVID-19’ in the population of COVID-19 vs. population. Leave-one-out analysis: each row represents a MR analysis of 25OHD concentration on COVID-19 using all instruments expect for the SNP listed on the y-axis. The point represents the beta with that SNP removed and the line represents 95% confidence interval. The summary data are reported by the COVID-19 host genetics initiative. COVID-19: Coronavirus disease 2019; MR: mendelian randomization; SNP, single-nucleotide polymorphism; 25OHD: 25-hydroxyitamin D; SD, standard deviation

**Figure 2.**
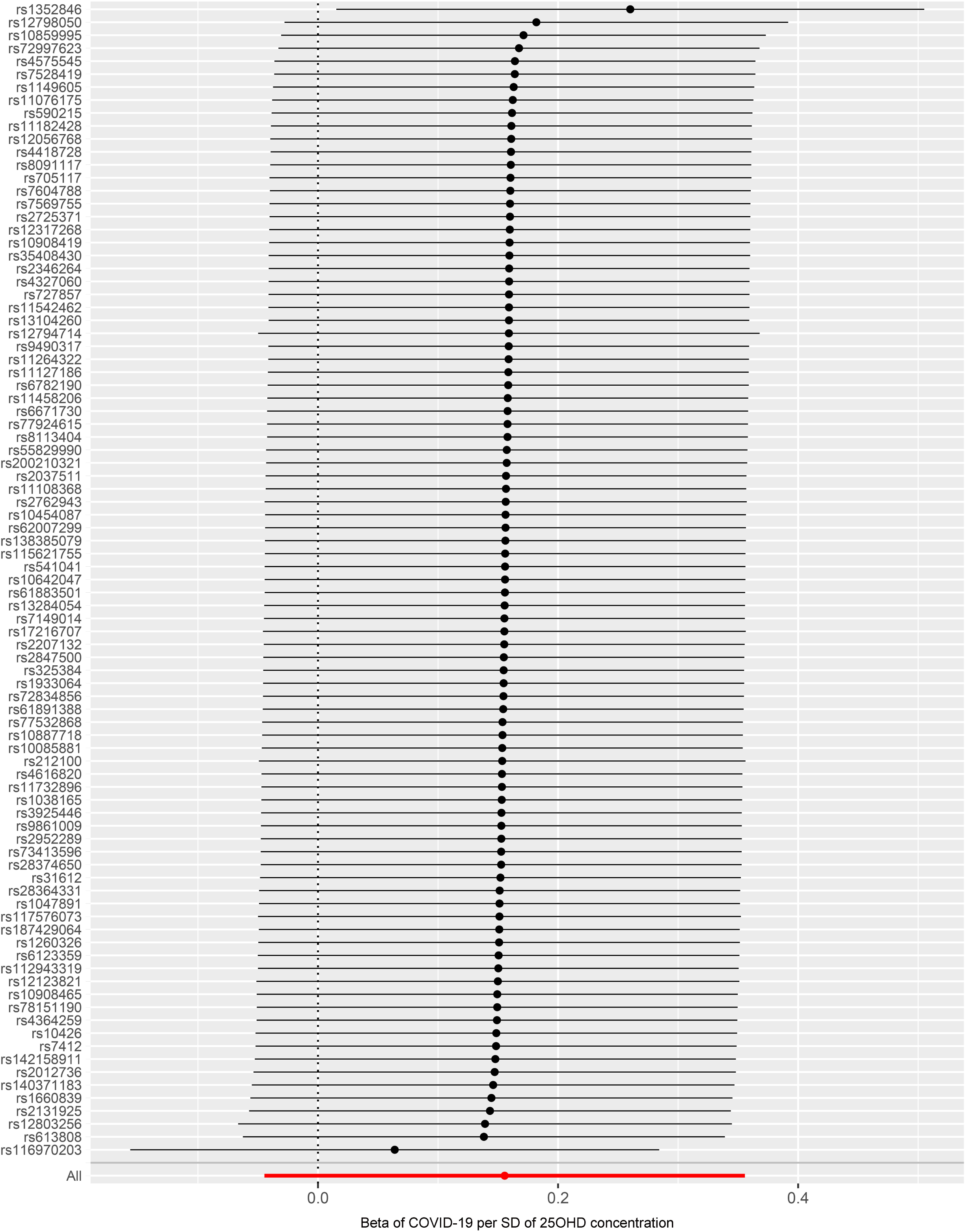
MR leave-one-out sensitivity analysis for ‘25OHD concentration’ on ‘severe COVID-19’ in the population COVID-19 vs. COVID-19 negative. Leave-one-out analysis: each row represents a MR analysis of 25OHD concentration on severe COVID-19 using all instruments expect for the SNP listed on the y-axis. The point represents the odds ratio with that SNP removed and the line represents 95% confidence interval. The summary data are reported by the COVID-19 host genetics initiative. COVID-19: Coronavirus disease 2019; MR: mendelian randomization; SNP, single-nucleotide polymorphism; 25OHD: 25-hydroxyitamin D; SD, standard deviation

The MR analysis showed no significant association of genetically instrumented 25OHD concentration with severe COVID-19 in the population of severe respiratory confirmed COVID-19 vs. population reported by two groups (OR, 0.889; 95% CI, 0.549-1.439; *P*=0.246; OR, 0.894; 95% CI, 0.587-1.363; *P*=0.603) (**Table 2**). The result in the weighted median, penalized weighted median, and MR-Egger regression methods were robust. There was limited evidence of heterogeneity and horizontal pleiotropy based on the Q test and MR-Egger intercept test. Sensitivity analyses using different instruments yielded similar findings, suggesting the robustness of the causal association (**Table 2, Figure 3-4, Figure S2**).

**Table 2.**
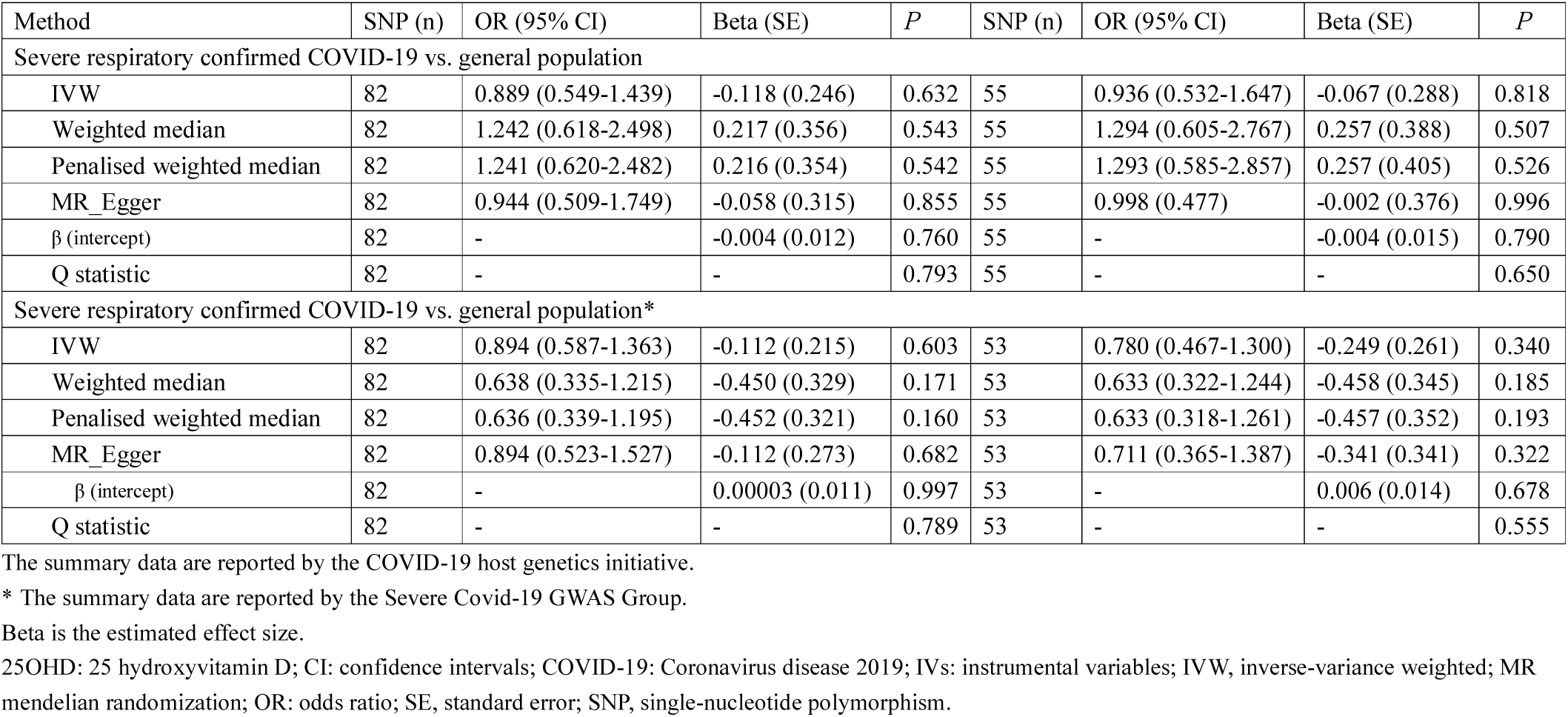
Causal association of 25OHD concentration with severe COVID-19.

**Figure 3.**
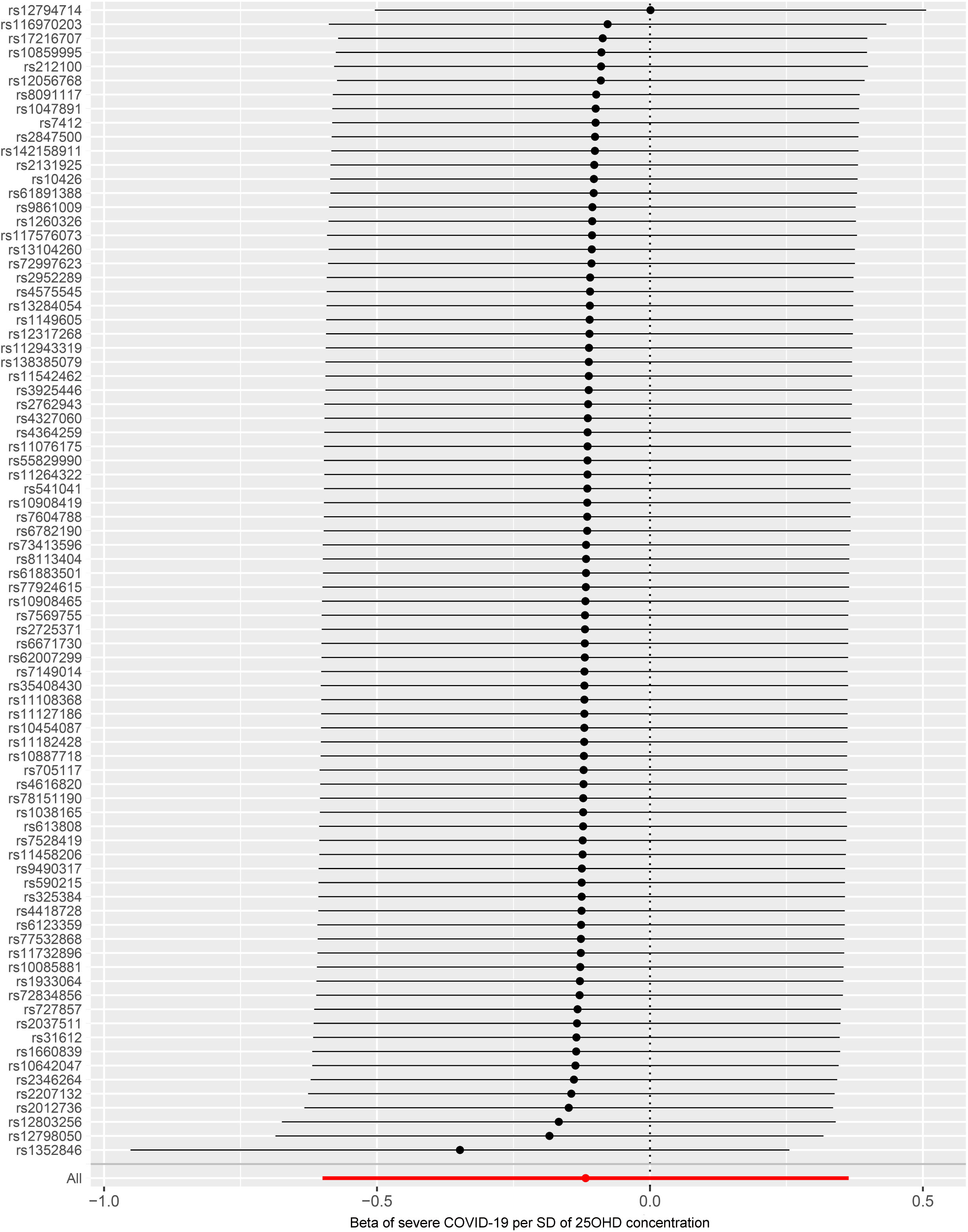
MR leave-one-out sensitivity analysis for ‘25OHD concentration’ on ‘severe COVID-19’ in the population of severe respiratory confirmed COVID-19 vs. population. Leave-one-out analysis: each row represents a MR analysis of 25OHD concentration on severe COVID-19 using all instruments expect for the SNP listed on the y-axis. The point represents the beta with that SNP removed and the line represents 95% confidence interval. The summary data are reported by the COVID-19 host genetics initiative. COVID-19: Coronavirus disease 2019; MR: mendelian randomization; SNP, single-nucleotide polymorphism; 25OHD: 25-hydroxyitamin D; SD, standard deviation

**Figure 4.**
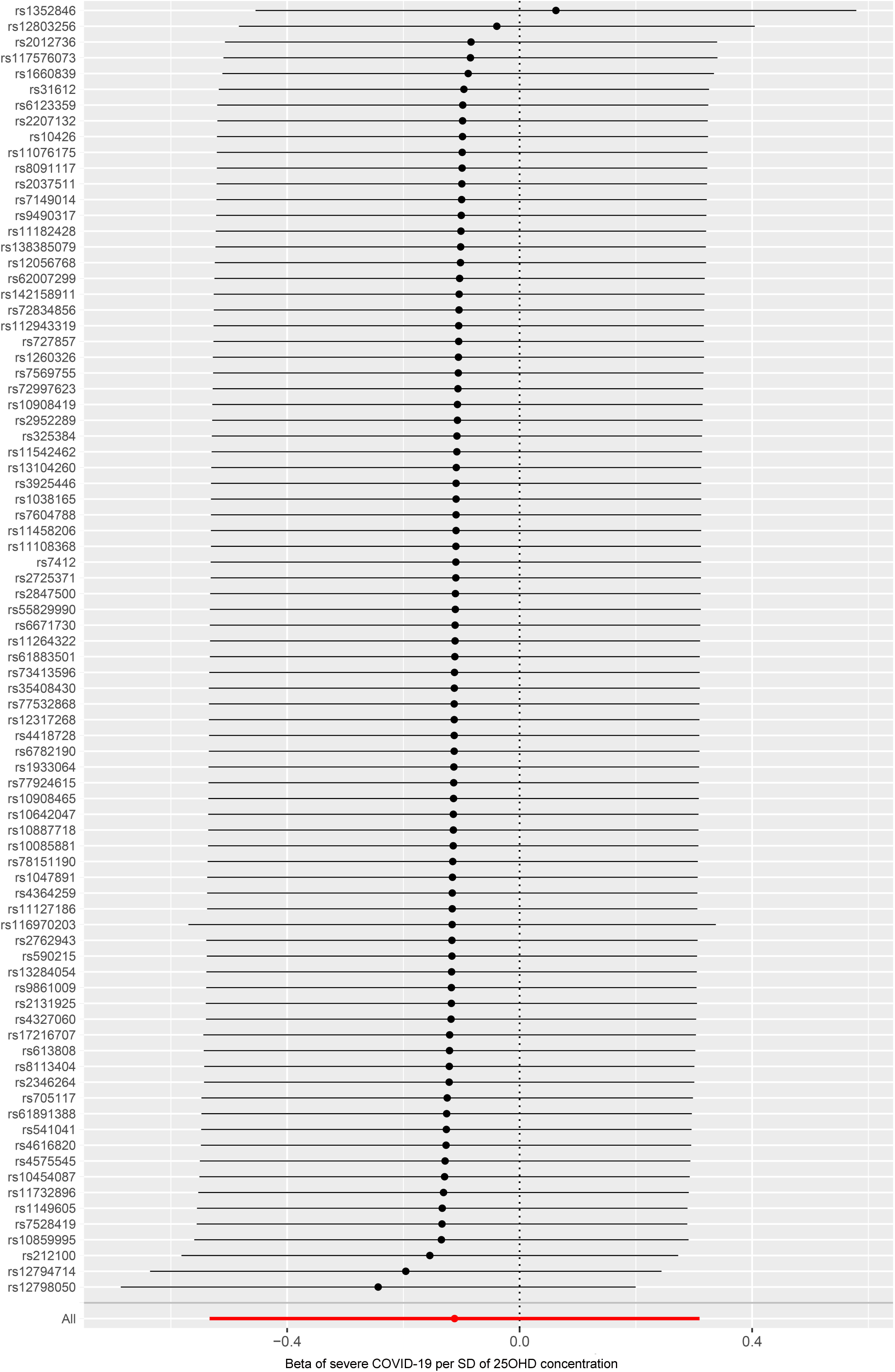
MR leave-one-out sensitivity analysis for ‘25OHD concentration’ on ‘severe COVID-19’ in the population of severe respiratory confirmed COVID-19 vs. population. Leave-one-out analysis: each row represents a MR analysis of 25OHD concentration on severe COVID-19 using all instruments expect for the SNP listed on the y-axis. The point represents the beta with that SNP removed and the line represents 95% confidence interval. The summary data are reported by the severe COVID-19 GWAS Group. COVID-19: Coronavirus disease 2019; MR: mendelian randomization; SNP, single-nucleotide polymorphism; 25OHD: 25-hydroxyitamin D; SD, standard deviation; GWAS, genome wide association study

## Discussion

In the current study, we performed an MR analysis to investigate the causal association of 25OHD concentration with the risk of COVID-19. Our results indicated that there might be no linear causal relationship of 25OHD concentration with COVID-19 susceptibility and severity.

An observational study based on the UK Biobank data claimed that no link between vitamin D concentrations and risk of COVID-19 infection either overall or separated ethnic groups ^16^. However, one study used causal inference analysis, supported the hypothesis that vitamin D plays a causal role in COVID-19 outcomes via modification of host responses to SARS-CoV-2^24^. In addition, there were also systematic reviews and meta-analyses to explore the association between vitamin D and COVID-19 ^9,10,25^ These studies provided a biological hypothesis and evolving epidemiological data supporting a role for vitamin D in COVID-19. But these results only based on the observational study design, which may be confounder bias. In our study, the SNPs associated with vitamin D as IVs were used to estimate the overall causal association of 25OHD concentration on COVID-19 susceptibility and severity, based on the MR design. The MR study could potentially avoid many biases and confounding issues existing in conventional observational studies and thus help to identify causally related risk factors. Using MR design, we found no evidence supporting that genetically predicted 25OHD concentration was significantly associated with COVID-19 susceptibility and severity.

There are some possible explanations for these negative findings. First, these null findings suggest that the associations of 25OHD concentration with COVID-19 susceptibility and severity could attribute from the reverse causation bias and confounder bias. Vitamin D from environment with across to adequate sunshine or diet was metabolized in the liver to 25OHD, which was used to determine a patient’s vitamin D status ^23,26^. Vitamin D deficiency may be common in COVID-19 patients, as a consequence of quarantined and reduced outdoor behavior. The observed association between 25OHD concentration with increased risk of COVID-19 could be confounded by outdoor behavior which may be corrected with the genetic liability to COVID-19 ^23^ Our findings suggested that COVID-19 susceptibility and severity are expected to decrease the prevalence of vitamin D deficiency, which are needed to be proved by more bi-directional MR studies. Second, it is also important to note that MR study considers the lifelong effect of genetic modification of COVID-19. However, the association between the vitamin D level and the risk of COVID-19 may be not fixed for a lifetime, but perform time-varying ^27^. The cross-sectional observational nature of all current MR studies limits the evaluation. The future MR studies incorporating follow-up data should be considered the effect of vitamin D level on COVID-19 and how genetic variants effects change with time may impact the interpretability and validity of their results. Third, as shown by previous studies, vitamin D supplementation only shows treatment effects among individuals with baseline 25OHD concentration of no more than 30 nmol/L, indicating that the relationship between the 25OHD concentration and the risk of diseases may be nonlinear ^28-30^. However, we noted that there is a linearity assumption in our Mendelian randomization analyses ^23^, then non-linear relationship could not be tested and might equate to the null hypothesis of no effect of the exposure on the outcome. Therefore, our results indicated that there might be no linear causal relationship of 25OHD concentration with COVID-19 susceptibility and severity.

The evidence of findings from MR studies sit at the interface between observational studies and RCTs ^31^. RCTs provide interventions for disease, while studies could therefore not be extrapolated for this purpose, but could rather be used to provide evidence of a casual relationship. It should be also focused on the effect of vitamin D supplementation on COVID-19. The previous findings suggest that vitamin D deficiency and treatment has a long-term effect on preventing overall mortality ^12,32^. In addition, future research should pay attention to not only the impact of vitamin D deficiency and treatment on the incidence of COVID-19, but also the impact of vitamin D deficiency and treatment on the COVID-19 mortality and lost life in COVID-19.

Some limitations should be noticed. It is important to note that the results of the MR analyses are based on numerous assumptions. First, we selected genetic variants as IVs based on the recent large-scale GWAS ^23^, which showed a strong association with 25OHD concentration; therefore, the bias of weak instrument might be less likely. Second, the genetic variants are not associated with measured and unmeasured confounders that influence both vitamin D and COVID-19. However, the unmeasured confounders or alternative causal pathways may be still affected our results because of the limitation of the method. Third, the existence of horizontal pleiotropy may distort MR results. In our study, there was limited evidence of heterogeneity and horizontal pleiotropy. In addition, the GWAS of the severe COVID-19 cases included small sample size, which might lead to small effect for the MR estimate and limit the IVs for COVID-19 for reverse MR analysis. The findings were based on European population, which made it difficult to represent the universal conclusions for other ethnic groups. Therefore, the future studies with larger sample size and more ethnic groups are needed to verify and explore the observed associations.

## Conclusion

Using 25OHD concentration-related SNPs as IVs from GWAS data, the MR analysis results indicated that there might be no linear causal relationship of 25OHD concentration with COVID-19 susceptibility and severity. In future, the bi-directional MR and non-linear MR study was needed to further prove these results. In addition, we should pay more attention to the randomization control trials about association between vitamin D treatment and the improvement of the COVID-19 in the long-term benefits.

## Data Availability

All data generated or analyzed during this study are included in this published article and its supplementary information files.

## Disclosure of Potential Conflicts of Interest

All authors have approved the manuscript and its submission. No potential conflicts of interest were disclosed by the authors.

## Funding/Support

The study was supported by grants from the National Natural Science Foundation of China (81872682 and 81773527), and the China-Australian Collaborative Grant (NSFC 81561128020-NHMRC APP1112767).

## Role of the Funder/Sponsor

The funding organization had no role in the design and conduct of the study; collection, management, analysis, and interpretation of the data; preparation, review, or approval of the manuscript; or decision to submit the manuscript for publication.

---

**Figure S1. MR leave-one-out sensitivity analysis for ‘25OHD concentration’ on ‘COVID-19’ in the population of COVID-19 vs. population**

Leave-one-out analysis: each row represents a MR analysis of 25OHD concentration on COVID-19 using all instruments expect for the SNP listed on the y-axis. The point represents the beta with that SNP removed and the line represents 95% confidence interval. The summary data are reported by the COVID-19 host genetics initiative. COVID-19: Coronavirus disease 2019; MR: mendelian randomization; SNP, single-nucleotide polymorphism; 25OHD: 25-hydroxyitamin D; SD, standard deviation

**Figure S2. MR leave-one-out sensitivity analysis for ‘25OHD concentration’ on ‘severe COVID-19’ in the population of COVID-19 vs. COVID-19 negative**

Leave-one-out analysis: each row represents a MR analysis of 25OHD concentration on severe COVID-19 using all instruments expect for the SNP listed on the y-axis. The point represents the odds ratio with that SNP removed and the line represents 95% confidence interval. The summary data are reported by the COVID-19 host genetics initiative. COVID-19: Coronavirus disease 2019; MR: mendelian randomization; SNP, single-nucleotide polymorphism; 25OHD: 25-hydroxyitamin D; SD, standard deviation

**Figure S3. MR leave-one-out sensitivity analysis for ‘25OHD concentration’ on ‘severe COVID-19’ in the population of severe respiratory confirmed COVID-19 vs. population**

Leave-one-out analysis: each row represents a MR analysis of 25OHD concentration on severe COVID-19 using all instruments expect for the SNP listed on the y-axis. The point represents the beta with that SNP removed and the line represents 95% confidence interval. The summary data are reported by the COVID-19 host genetics initiative.

COVID-19: Coronavirus disease 2019; MR: mendelian randomization; SNP, single-nucleotide polymorphism; 25OHD: 25-hydroxyitamin D; SD, standard deviation

**Figure S4. MR leave-one-out sensitivity analysis for ‘25OHD concentration’ on ‘severe COVID-19’ in the population of severe respiratory confirmed COVID-19 vs. population**

Leave-one-out analysis: each row represents a MR analysis of 25OHD concentration on severe COVID-19 using all instruments expect for the SNP listed on the y-axis. The point represents the beta with that SNP removed and the line represents 95% confidence interval. The summary data are reported by the severe COVID-19 GWAS Group.

COVID-19: Coronavirus disease 2019; MR: mendelian randomization; SNP, single-nucleotide polymorphism; 25OHD: 25-hydroxyitamin D; SD, standard deviation; GWAS, genome wide association study

## Notes

### Competing Interest Statement

The authors have declared no competing interest.

### Author Declarations

We used the summarized data. Ethical information is not available.

### Summary of Updates

We used another summarized data for severe respiratory confirmed COVID-19 reported by from the Severe Covid-19 GWAS Group for further analysis.

